# Predictive Time Series Modeling of Malaria Incidence in India: A 20-Year Retrospective and 5-Year Forecast

**DOI:** 10.1101/2024.09.26.24314440

**Authors:** Ami Soni

## Abstract

**Background:** Malaria poses a significant health challenge in India, with notable contributions to the global malaria burden. This study aims to analyze trends in malaria incidence in India over the past two decades, evaluating the effectiveness of public health interventions.

**Objective:** The objective of this study is to analyze trends in malaria incidence in India over the past decades, evaluate the effectiveness of public health interventions, and project future trends in order to support the country’s goal of malaria elimination by 2030.

**Methods:** A comprehensive 20-year dataset was utilized to conduct time series analyses, including linear regression and ARIMA modeling. The linear regression model assessed the trend in malaria incidence per 1,000 population at risk, while the ARIMA model was used for forecasting future trends. Residuals were evaluated for adequacy to ensure model reliability.

**Results:** The linear regression analysis revealed a significant annual decrease in malaria incidence by approximately 0.92 units (p < 0.001), explaining 90.77% of the variability in the data. The ARIMA model forecasts indicate a continued decline, projecting negative incidence values by 2026 and 2027, despite some residual autocorrelation suggesting further model refinement may be necessary.

**Discussion:** These findings highlight the effectiveness of current public health strategies and the importance of ongoing monitoring to address remaining challenges. The projected downward trend aligns with India’s goal of malaria elimination by 2030, reinforcing the need for sustained interventions in high-burden areas.

**Conclusion:** This study underscores the positive impact of public health initiatives on malaria incidence in India, while emphasizing the necessity for continuous research and adaptive strategies to achieve the ambitious target of malaria eradication by 2030.

## Introduction

Malaria is a potentially life-threatening illness caused by several parasitic species, namely Plasmodium vivax, Plasmodium falciparum, Plasmodium malariae, and Plasmodium ovale, and is transmitted through the bite of infected female Anopheles mosquitoes (1). In 2022, approximately 249 million malaria cases were reported globally across 85 endemic countries, reflecting an increase of 5 million cases compared to the previous year (2). Within the WHO South-East Asia Region, nine countries were identified as malaria-endemic, contributing to 5.2 million cases, which represented 2% of the global malaria burden (1). India’s significant malaria case load places it among the eleven countries with the highest burden which collectively account for 70% of the global incidence of malaria as seen in Figure 1, with the remaining ten countries located in Africa. India accounting for roughly 65.7% of all malaria cases within the south-eastern region, with nearly 46% attributed to P. vivax. Together, India and Indonesia were responsible for 94% of malaria-related fatalities in the south-east region (4).

**Figure 1.**
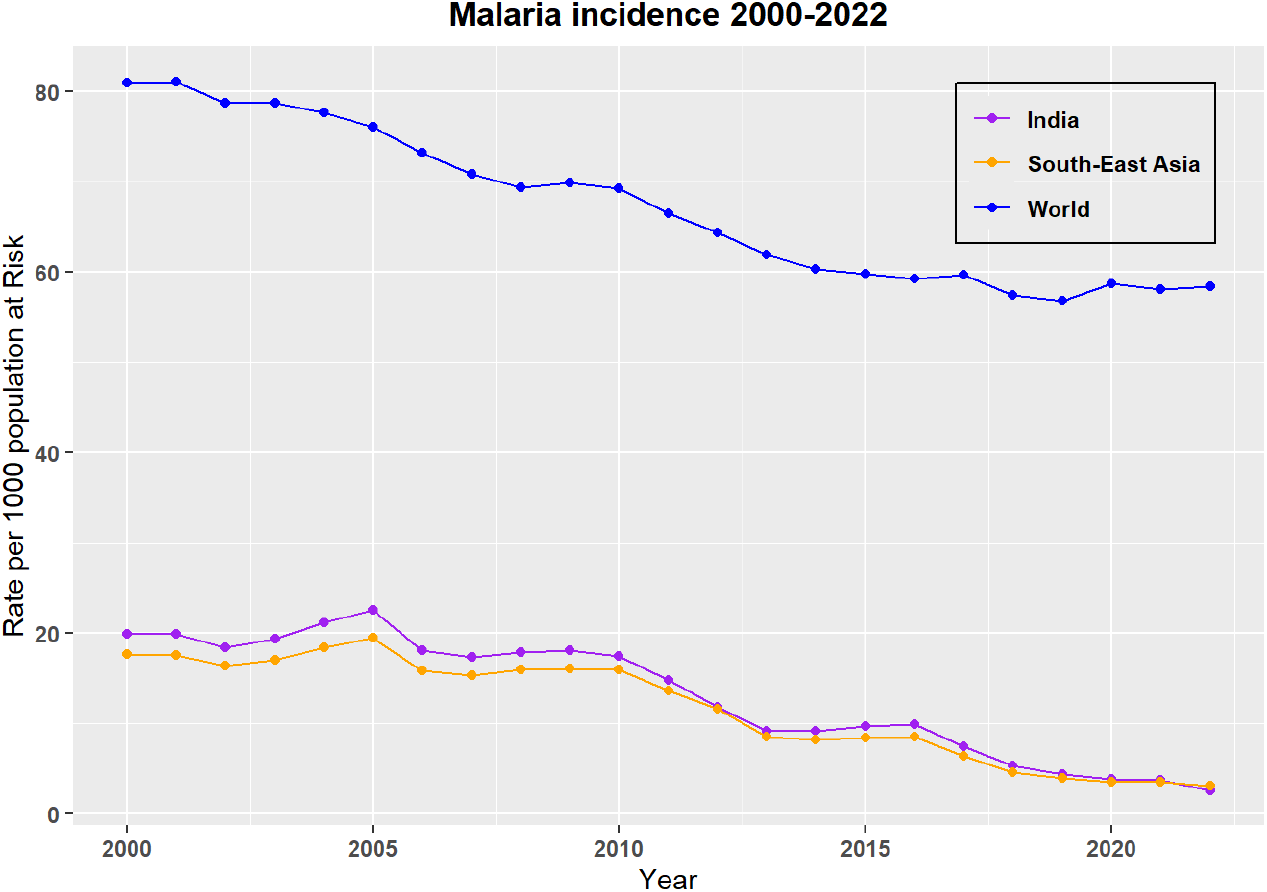
Malaria incidence in World, South-East Asia and India (2000-2022)

To combat this public health challenge, the World Health Assembly adopted the “Global Technical Strategy for Malaria 2016–2030,” which guides malaria elimination initiatives. The WHO Global Malaria Programme (GMP) is tasked with co-ordinating international efforts focused on controlling and ultimately eliminating malaria. Aligned with this strategy in February 2016 with the assistance of WHO, the Government of India launched the National Framework for Malaria Elimination 2016–2030, followed by the National Strategic Plan for Malaria Elimination 2017–2022 in July 2017. India’s goal is to attain malaria - free status by 2027 and achieve complete eradication by 2030. Under the NSP 2017-2022, the WHO has supported the acceleration of malaria elimination initiative across several states. In 2018 WHO launched High Burden and High Impact(HBHI) initiative which seeks to strengthen malaria control efforts in high burden nation. Follwoing WHO, in july 2019, India introduced the High Burden of High Impact (HBHI) strategy in four highly endemic states: West Bengal, Jharkhand, Chhattisgarh and Madhya Pradesh.Despite these efforts, challenges remain due to the extensive diversity of ecosystems in India, with approximately 80% of the malaria burden concentrated in only ten of the 28 states, six of which are located in the northeastern part of the country (3,4).

## Methods

This study employs a comprehensive 20-year dataset to analyze malaria incidences in India, utilizing time series analysis as a crucial approach for understanding and forecasting trends in endemic regions. A thorough examination of historical data was conducted to identify key components, including underlying trends and irregular fluctuations. Given the annual frequency of the data and the absence of a clear seasonal component, traditional decomposition methods such as `decompose()` and `stl()` were deemed unsuitable, as these approaches typically require larger datasets to effectively capture seasonal patterns.

Consequently, a linear regression model was fitted to the time series to analyze the trend, with the rate of malaria incidence per 1,000 population at risk as the dependent variable and time as the independent variable. The results were visualized to illustrate the time series data alongside the fitted trend line, as depicted in Figure 3.

**Figure 2.**
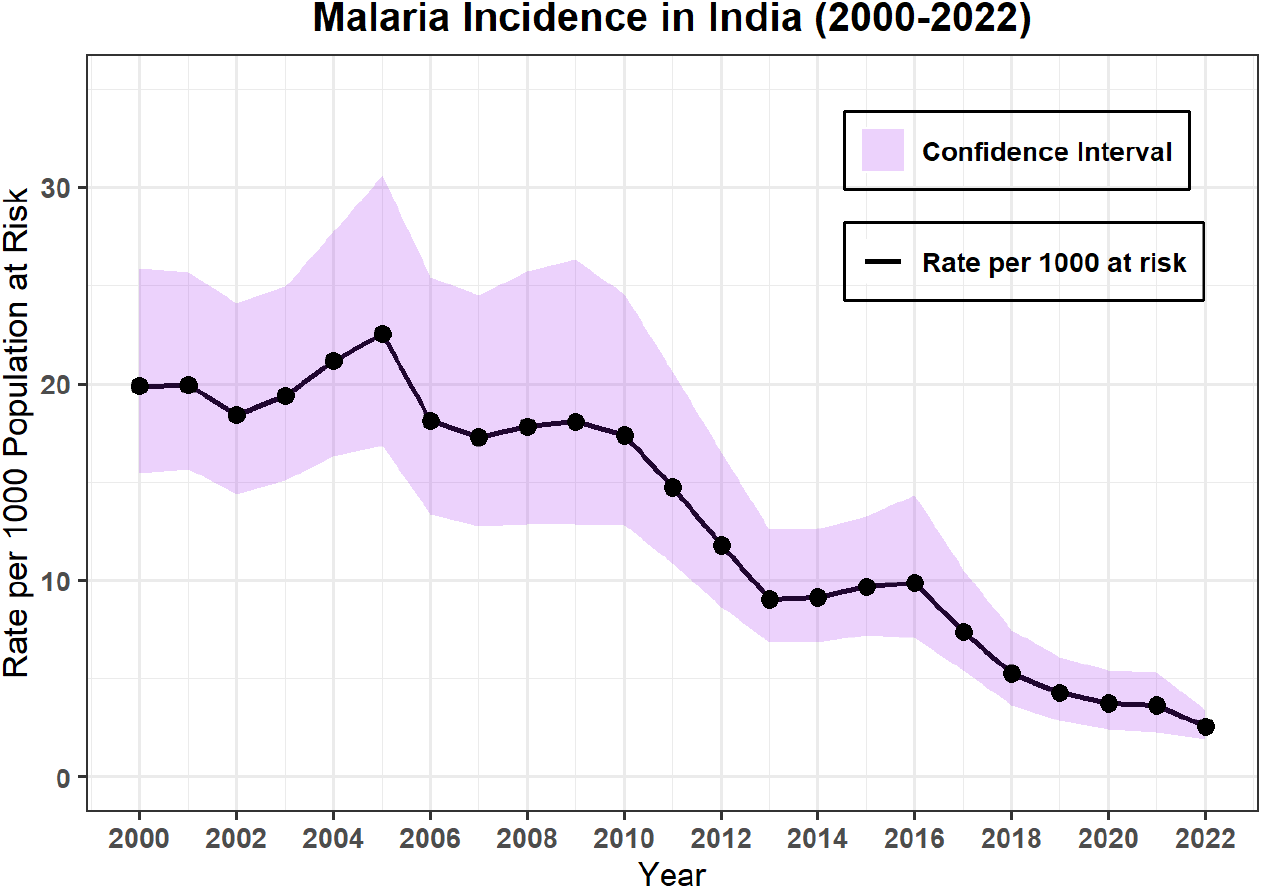
Malaria incidence of India with confidence internval.

**Figure 3.**
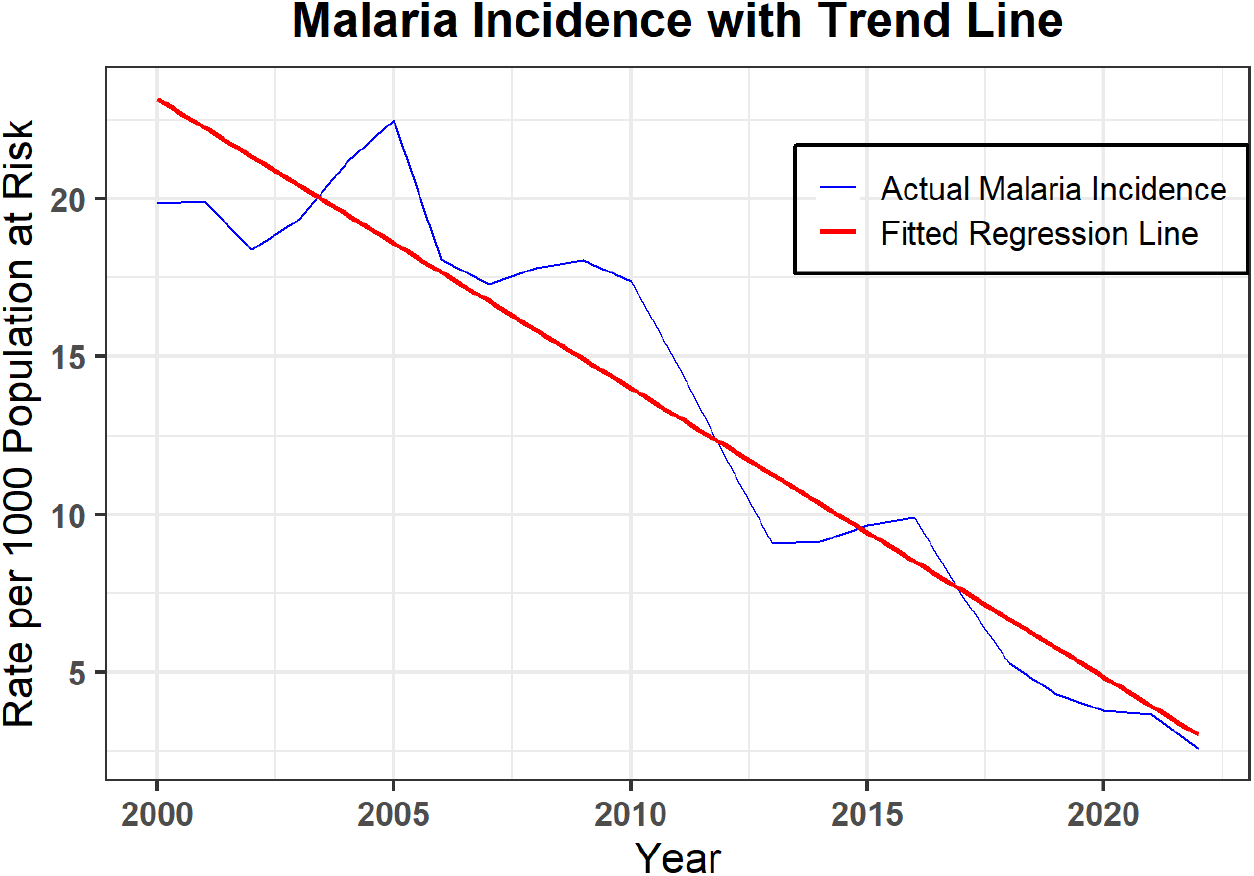
Linear regression model to the time series analysis.

**Figure 4.**
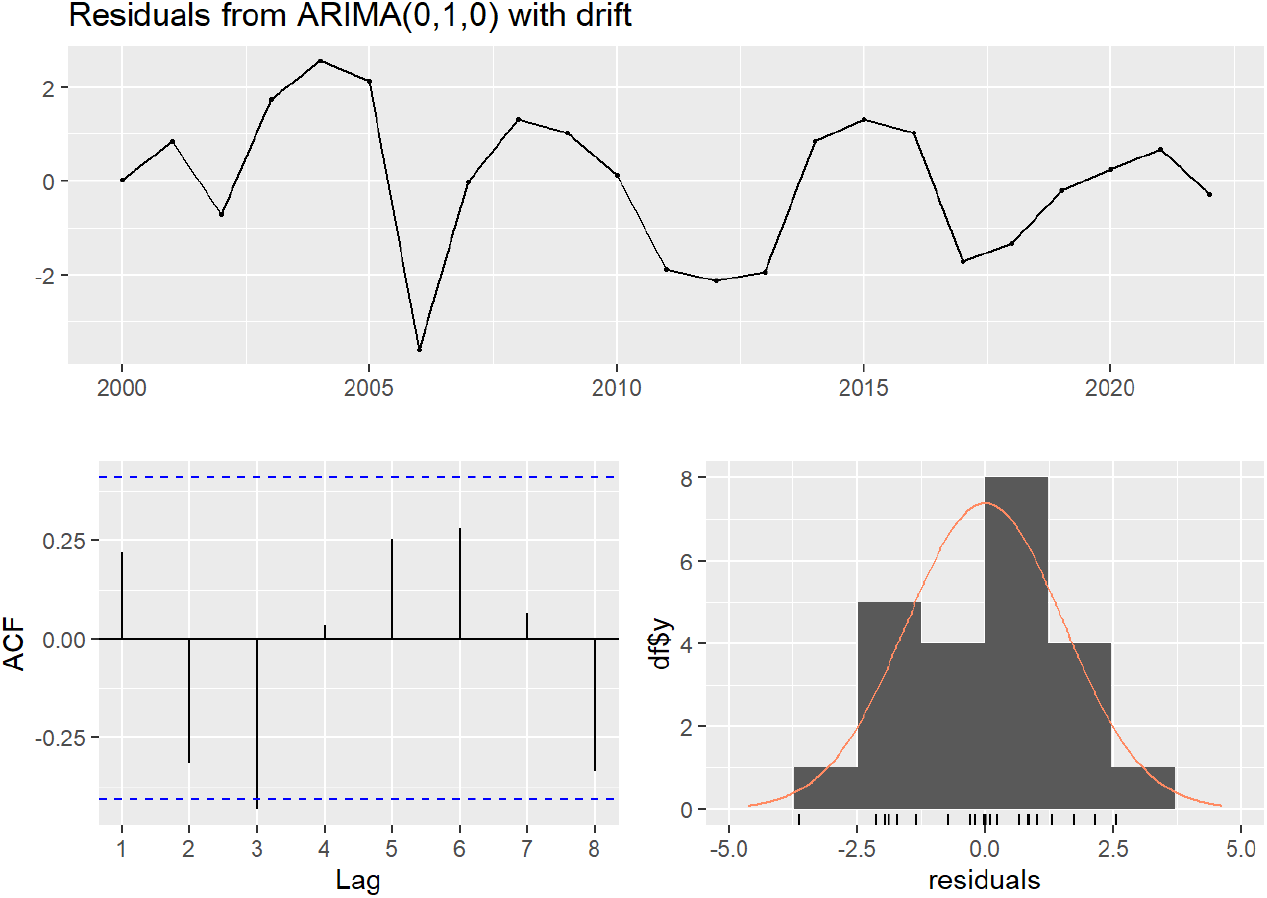
Residuals with Arima Model (0,1,0)

Following the linear regression analysis, ARIMA modeling techniques were employed using the `auto.arima()` function in R, which automates the selection of appropriate ARIMA parameters. The ARIMA model was fitted to the malaria incidence time series data to capture any potential autocorrelations and temporal dependencies that may not have been addressed by the linear regression model.

To project future malaria incidence, the `forecast(fit, h = 5)` function was utilized, allowing for the generation of forecasts for the next five years, which are presented alongside their confidence intervals in Figure 6. Throughout this analysis, a rigorous data cleaning process was conducted to ensure the integrity and reliability of the results.

The residuals of both the linear regression and ARIMA models were evaluated for adequacy. The `checkresiduals(fit)` function was used to assess the ARIMA model’s residuals, ensuring that no patterns remained unexplained. Additionally, the Autocorrelation Function (ACF) and Partial Autocorrelation Function (PACF) were employed to generate plots that further analyze the residuals, confirming that the models adequately captured the time-dependent structure of the data.

By leveraging these advanced analytical methods, this study aims to elucidate trends in malaria incidence in India, thereby supporting data-driven public health strategies designed to achieve malaria elimination by 2030. A comprehensive understanding of these trends is critical for informing targeted interventions in high-burden areas and optimizing resource allocation effectively.

## Results

### Linear Regression Analysis

The linear regression analysis revealed significant findings:

**Table 1a.** Coefficients of the Trend model.

| Term | Estimate | Std_Error | t_value | p_value |
| --- | --- | --- | --- | --- |
| (Intercept) | 1858.76385 | 128.40385 | 14.48 | 2.13e-12 |
| Time | -0.91779 | 0.06385 | -14.37 | 2.44e-12 |

**Table 1b.** Residuals & Summary Statistics of the Trend Model.

| Statistic | Value |
| --- | --- |
| **Residuals** |  |
| Min | -3.3219 |
| 1Q | -1.3004 |
| Median | -0.2758 |
| 3Q | 1.5122 |
| Max | 3.9144 |
| **Summary Statistic** |  |
| Residual Standard Error | 2.031 |
| Multiple R-squared | 0.9077 |
| Adjusted R-squared | 0.9033 |
| F-statistic | 206.6 |
| p-value | 2.441e-12 |

### Coefficients

- *Intercept:*
  - The estimated baseline incidence rate at the start of the time series was 1858.76 (p < 0.001), serving as a reference point despite not correspinding to a real value due to data starting in 2000.
- *Time:*
  - The coefficient of -0.91779 indicates a significant annual decrease in malaria incidence by approximately 0.92 units (p < 0.001), highlighting a statistically significant downward trend.

### Residuals

The residuals range form -3.32 to 3.91, suggesting that while the model fits well, there are some unexplained variations.

### Model Fit

- *Residual Standard Error:*
  - At 2.031, this value indicates a relatively good fit of the model.
- *Multiple R-squared:*
  - The value 0.9077 suggest that the model explains about 90.77% of the variability in malaria incidence, demonstrating a strong fit.
- *Adjusted R-squared:*
  - At 0.9033, this value confirms the model’s robustness while accounting for the number of predictors.
- *F-statistics and P-value:*
  - The value 206.6 of F-statitsics, with a *P-value* of 2.441e-12, indicates that the model is statistically significant overall.

### Forecasting with ARIMA

The application of the ARIMA model yielded the following findings:

### Residuals check

The residuals were analyzed, yielding the following results. Table.2 Residuals from ARIMA(0,1,0) with drift.

**Table 2:** Residuals from ARIMA (0,1,0) with drift.

| Statistic | Value |
| --- | --- |
| Q* | 11.447 |
| Degrees of Freedom | 5 |
| p-value | 0.04321 |
| Model df | 0 |
| Total lags used | 5 |

### Q-statistics

The Q-statistics of 11.447 and p-value of 0.04321 indicates that there is some remaining autocorrelation in the residuals, suggesting that the model model may not fully capture the underlying structure of the data.

### ACF & PACF Analysis

The ACF & PACF plots provided disgnostics insights into the model’s fit, as seen in Figure 5. The ACF plot revealed minimal autocorrelation among the residuals across various time lags, with most points falling within the confidence interval. However, minor spikes, particularly at lag 1, suggest some residual autocorrelation though the overall effect is likely minimal. The PACF plot confirmed that most partial autocorrelation values lie within the confidence bounds, indicating no significant correlation remains among the residuals after accounting for previous lags. A small spike at lag 1 suggests potential short-term autocorrelation.

**Figure 5.**
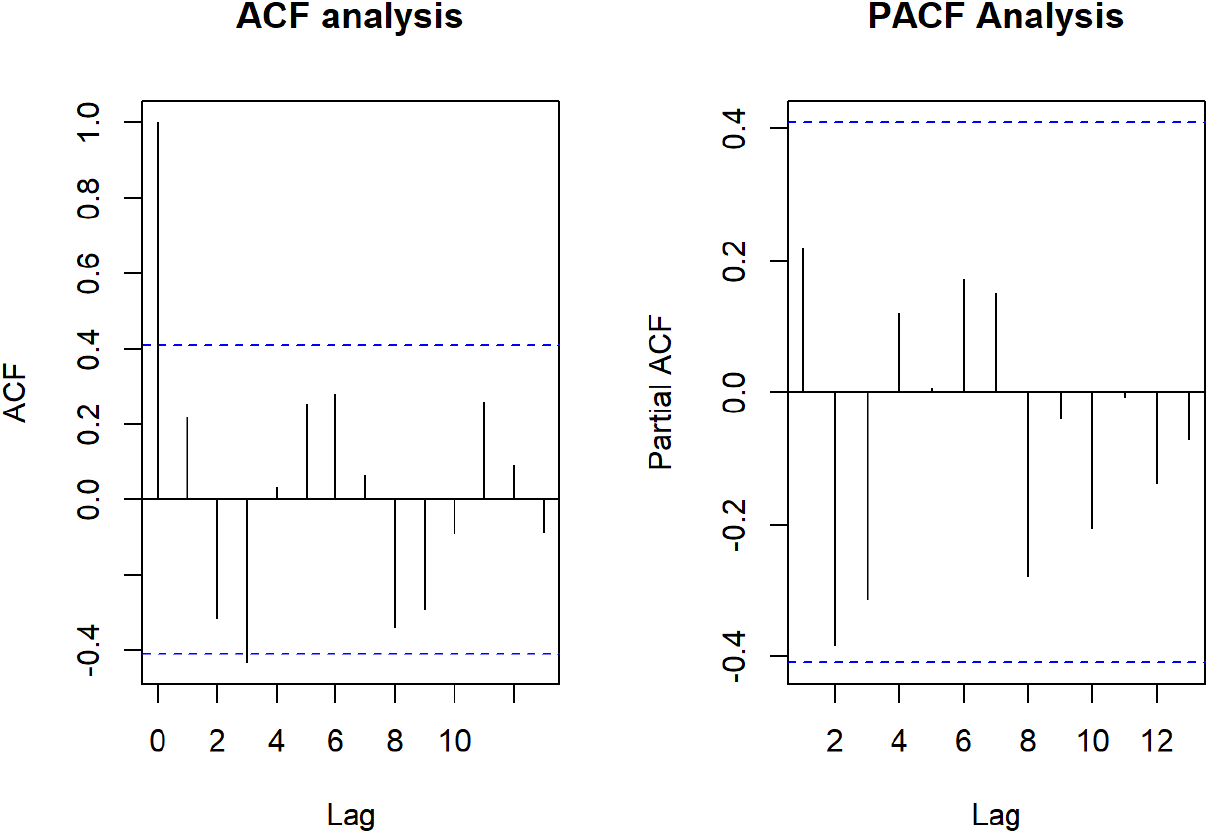
ACF and PACF Analysis.

### Model Fit

Overall, the model effectively captured the underlying structure of the malaria incidence data, with residuals behaving largely like white noise. However, the minor autocorrelation at lag 1 suggests opportunities for model refinement.

### Forecast for the Next Five Years

The forecasts for the upcoming years are illustrated in Figure 6 and summarized in the following Table 3. The projections indicate an estimated incidence of approximately 1.77 cases in 2023, declining to 0.99 cases in 2024, with further decreases projected for subsequent years, ultimately suggesting negative incidence values by 2026 and 2027. It is important to note that the wide confidence intervals associated with these forecasts reflect variability in the predictions, underscoring the inherent uncertainty in forecasting future malaria incidences.

**Figure 6.**
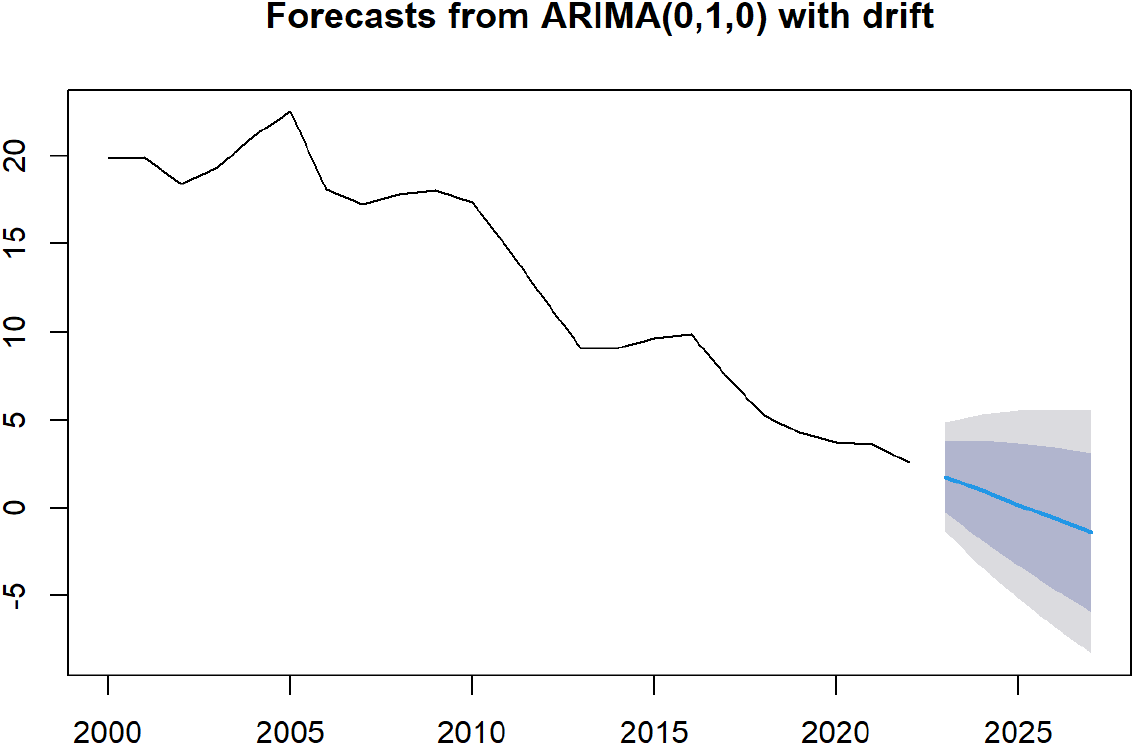
Forecast of Malaria Incidence in India for next 5 years.

**Table 3:** Forecast of Malaria Incidence in India for next five years.

| Year | Point Forecast | Lo 80 | Hi 80 | Lo 95 | Hi 95 |
| --- | --- | --- | --- | --- | --- |
| 2023 | 1.7729418 | -0.2458667 | 3.791750 | -1.314559 | 4.860443 |
| 2024 | 0.9863136 | -1.8687128 | 3.841340 | -3.380072 | 5.352700 |
| 2025 | 0.1996855 | -3.2969935 | 3.696364 | -5.148023 | 5.547394 |
| 2026 | -0.5869427 | -4.6245598 | 3.450674 | -6.761945 | 5.588060 |
| 2027 | -1.3735709 | -5.8877640 | 3.140622 | -8.277434 | 5.530292 |

## Discussion

### LINEAR Regressions

The findings from the linear regression analysis provide valuable insights into the trends of malaria incidence rates in recent years. The observed annual decrease of approximately 0.92 units strongly suggests that the public health interventions implemented are making a positive impact on malaria control. This downward trend is not only encouraging but also aligns with global efforts to reduce malaria transmission, reinforcing the effectiveness of targeted health strategies.

Despite the model’s ability to explains a substantial portion of the variability in malaria incidence, the residuals ranging from -3.32 to 3.91 indicate that there are still unexplained variations. These residuals may be influenced by factors such as local environmental conditions, socio-economic disparities, or variations in health service accessibility, which were not accounted for in the model. This highlights the complexity of malaria dynamics and the need for comprehensive approaches that address these underlying factors.

The high multiple R-squared value demonstrates the model’s efficacy in explaining the observed data, while the adjusted R-squared confirms its robustness after considering the number of predictors. The strong F-statistic and its corresponding p-value reinforces the overall significance of the model, indicating that the predictors employed are indeed relevant to understanding malaria trends.

### ARIMA Model & Forecast

Transitioning to the ARIMA modeling and forecasting analysis, the results provides further insights into malaria incidence trends in India. The projected decline in malaria cases aligns with the positive impact of ongoing public health interventions. The model’s effectiveness is evidenced by minimal autocorrelation observed in the ACF and PACF plots, suggesting that the fitted model adequately captures the temporal dynamics of malaria incidence.

However, the presence of statistically significant autocorrelation in the residuals indicates that some patterns remain unaccounted for. This suggests opportunities for refinement in the model, particularly concerning the minor autocorrelation observed at lag 1. Future model iterations could explore alternative specifications or additional predictors to enhance fit and predictive accuracy.

The forecasting results reveals a significant reduction in malaria incidence, with projections indicating a decline to negative values by 2026 and 2027. While these findings are encouraging, they necessitates cautious interpretation due to the wide confidence intervals that reflect inherent uncertainty. Continuous monitoring of malaria incidence trends and adaptive management strategies will be crucial for sustaining this positive trajectory and achieving malaria elimination goals by 2030.

Overall, the results underscore a significant downward trend in malaria incidence, aligning with India’s public health initiatives aimed at malaria elimination. The study’s findings suggest that current interventions are effective, yet ongoing efforts remain essential, particulary in high-burden regions. The data-driven approaches utilized in this analysis are vital for understanding and forecasting malaria trends, thereby supporting public health strategies aimed at achieving malaria eradication by 2030.

As India strives for malaria-free status by 2027, these findings emphasize the critical need for sustained public health interventions to overcome remaining challenges. The effective capture of historical patterns and projection of future trends through the ARIMA model highlighting the importance of ongoing monitoring and evaluation in the fight against malaria.

## Conclusion

This study provides a comprehensive analysis of malaria incidence in India over the past two decades, employing predictive time series modeling techniques to uncover significant trends. The findings reveal a marked downward trajectory in malaria cases, which underscores the positive impact of ongoing public health interventions. However, achieveing the ambitious goal of malaria eradication by 2030 will require Continued monitoring and the implementation of targeted strategies.

The analysis not only highlights the effectiveness of current malaria control efforts but also emphasizes the importance of ongoing research. Identifying and addressing the remaining factors contributing to variability in malaria incidence is essential for refining interventions. A nuanced understanding of these dynamics will be critical in enhancing the effectiveness of public health strategies and ensuring resource allocation is optimized.

In summary, this analysis reaffirms the efficacy of current public health initiatives in reducing malaria transmission while underscoring the necessity for continous research and model refinement. These efforts will be pivotal in sustaining progress towards malaria elimination and realizing a malaria-free future for India by 2030.

## Data Availability

The dataset used in this study are available from World Health Oraganization website.(2)

## Source Code

The source code for this study is available at GitHub/Dr-A-Soni. (https://github.com/Dr-A-Soni/Malaria_Incidence_Trends)

